# Efficient Edge-AI Models for Robust ECG Abnormality Detection on Resource-Constrained Hardware

**DOI:** 10.1101/2023.08.31.23294925

**Authors:** Zhaojing Huang, Luis Fernando Herbozo Contreras, Wing Hang Leung, Leping Yu, Nhan Duy Truong, Armin Nikpour, Omid Kavehei

## Abstract

This study introduces two models, CLTC and CCfC, designed for abnormality identification using ECG data. Trained on the TNMG subset dataset, both models were evaluated for their performance, generative capacity, and resilience. They demonstrated comparable results in terms of F1 scores and AUROC values. The CCfC model achieved slightly higher accuracy, while the CLTC model showed better handling of empty channels. Remarkably, the models were successfully deployed on a resource-constrained microcontroller, proving their suitability for edge device applications. Generalization capabilities were confirmed through the evaluation of the CPSC dataset. The models’ efficient resource utilization, occupying 70.6% of total storage and 9.4% of flash memory, makes them promising candidates for real-world healthcare applications. Overall, this research advances abnormality identification in ECG data, contributing to the progress of AI in healthcare.

## 1 Introduction

Various technologies have been developed to monitor heart activities, and among them, the Electrocardiogram (ECG) has gained widespread use due to its non-invasiveness and cost-effectiveness. In clinical settings, the 12-lead ECG is currently considered the standard for measuring cardiac electrical activity. This technique entails positioning 12 leads, consisting of six limb leads and six chest leads, to capture heart activities from both the vertical and horizontal planes [1].

This study presents novel neural circuit policies (NCP) empowered models aimed at assisting clinicians in detecting the specific location where abnormality occurs. The models, as opposed to traditional recurrent neural networks such as Long Short-Term Memory (LSTM), offer the advantage of mitigating the negative effects of learning long-term dependencies on specific tasks [2]. According to the study by Mathias, the NCP model demonstrates superior computational capabilities for neurons compared to contemporary deep models [2]. Unlike common deep neural network models, which rely heavily on unpolluted input data, the NCP model exhibits higher tolerance to transient disturbances that are common in real-world conditions [2]. Additionally, the NCP model’s compact and sparse network architecture eases the interpretation process [2]. Furthermore, the model requires low memory usage, making it suitable for deployment on microcontrollers. Another advantage is that the NCP model requires only a small number of neurons. However, it has been observed that this enhancement comes at the cost of reduced accuracy performance. The current model achieved an F1 accuracy of 0.82. The vision of the work, depicted in Fig. 1, can be extended to micro-controllers, allowing its integration into wearable devices. Indeed, the efficient resource utilization of the models is a noteworthy aspect, as they occupy a mere 240 KB, which is approximately 70.6% of the total RAM available on the STM32F746G microcontroller (equipped with 340 KB of RAM). Moreover, their utilization of approximately 96 KB of flash memory storage accounts for about 9.4% of the total flash memory available on the board, which boasts 1 MB of flash memory. This optimized use of resources renders the models highly promising candidates for real-world healthcare applications, where limitations in storage and memory capacity are critical considerations. Furthermore, the current on-chip model demonstrates reasonable power consumption, measuring at 137.4 mW. This level of power consumption is particularly crucial for ensuring that the models can operate efficiently in battery-powered devices without rapidly draining the power source. The power efficiency of these models enhances their suitability for various healthcare applications, enabling continuous and reliable operation while conserving energy resources.

**Figure 1:**
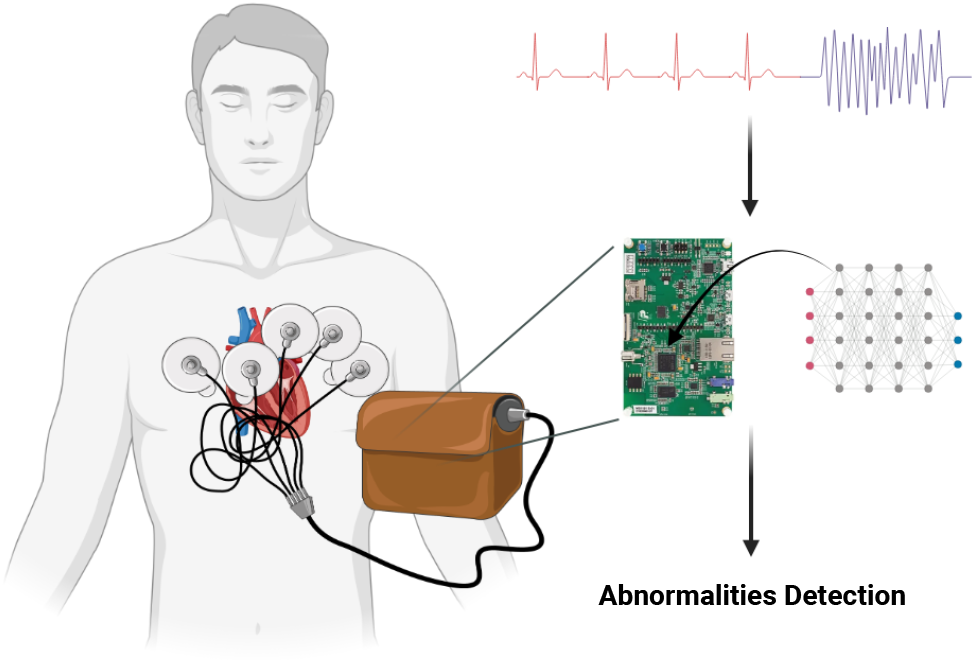
The model’s vision revolves around its application in wearable devices, where it plays a crucial role in detecting abnormalities and aiding in their identification.

### 1.1 Background

In the field of abnormality detection, several notable models have been developed. Georgios proposed a Hybrid CNN-LSTM Network, achieving a sensitivity of 97.87% and specificity of 99.29% for classifying a specific heartbeat type using ECG data [3]. Varun Gupta introduced a model that utilized fractional wavelet transform for pre-processing, Yule-Walker Autoregressive Analysis for feature extraction, and Principal Component Analysis for detection. This model achieved remarkable results, including a mean square error of 0.1656%, detection accuracy of 99.89%, and an output signal-to-noise ratio of 25.25 dB [4]. Tsai-Min Chen developed a model consisting of 5 CNN blocks, a bidirectional RNN layer, attention layer, and dense layer. This model achieved an F1 score of 0.84 in the detection of different types of ECG [5]. Jing-Shan Huang proposed a fast compression residual convolutional neural network-based model for ECG classification, which achieved an average accuracy of 98.79% [6]. Huang and Contreras developed an shallow S4D model, which demonstrated robust F1 score of 0.81 and high robustness for the input data [7].

For the NCP model, there is also several notable models have been developed. Mathias Lechner’s NCP model, originally developed for car driving applications using input from cameras, exhibits a distinct focus on the road’s horizon compared to conventional CNN models. While CNN models often prioritize roadside features and overlook the road itself, the NCP model demonstrates a different approach, placing greater emphasis on the road’s horizon [2]. This unique characteristic allows the NCP model to capture and learn global driving features effectively. This is reflected in the high variance explained by the first principal component (PC1), which reaches 92% [2]. The ability of the NCP model to concisely learn global driving features can contribute to improved understanding and decision-making in car driving applications. Ramin Hasani made advancements in Liquid Time-constant Networks by developing closed-form continuous-time neural networks, which exhibit improved computational speed [8].

## 2 Prerequisite

### 2.1 Liquid Time-Constant (LTC) Networks

The equation 1 describes LTC neurons, where *τ*_*i*_ represents the time constant of neuron *i*, determined by the ratio of its membrane capacitance 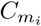 to leakage conductance 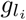 [2]. The activation function *σ*_*i*_(*x*_*j*_(*t*)) captures the behavior of neuron *i*, using the input *x*_*j*_ from neuron *j* and parameters *γ*_*ij*_ and *μ*_*ij*_ to shape a sigmoid curve output between 0 and 1 [2]. The synaptic weight *w*_*ij*_ reflects the strength of the connection from neuron *i* to *j*, while 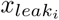 represents the resting potential of neuron *i* [2]. Additionally, *E*_*ij*_ defines the polarity of the synapse, determining its excitatory or inhibitory nature [2].

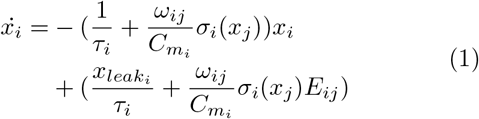

### 2.2 Neural circuit policies (NCP)

NCP model is an end-to-end learning system with several convolution layers [2]. In the neural circuit policies (NCP) model, four neural layers are incorporated: sensory neurons (Ns), interneurons (Ni), command neurons (Nc), and motor neurons (Nm). Between each consecutive layer, a specific number of synapses are inserted to facilitate the flow of information [2]. Firstly, between two consecutive layers, source neurons transmit information to target neurons through a specified number of synapses (nso-t), where the synapse distribution follows a Bernoulli distribution with a probability of p2 [2]. Secondly, for target neurons that do not have any synapses, additional synapses (mso-t) are introduced from source neurons, with the number of synapses determined by a Binomial distribution with a probability of p3 [2]. These synapses are randomly selected from a pool of source neurons. Thirdly, command neurons exhibit recurrent connections, where synapses (lso-t) are established to target command neurons, selected from a Binomial distribution with a probability of p4 [2]. Overall, these mechanisms contribute to the information flow and computational dynamics within the NCP model [2].

The given equation 2 illustrates the semi-implicit Euler technique applied to the NCP model, where *I*_*in*_ denotes the collection of neurons that serve as inputs to neuron *i*:

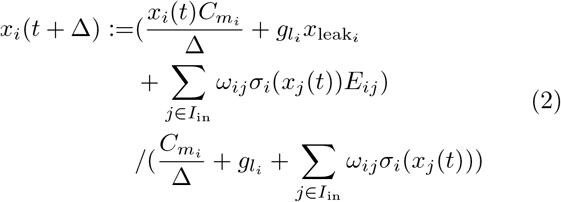

### 2.3 Closed-form Continuous-time (CfC) Neural Networks

The CfC model is built up by input perception module, LTC module and outputs[9]. A notable characteristic of Closed-form Control (CfC) neural networks is that they do not rely on numerical Ordinary Differential Equation (ODE) solvers to generate their temporal rollouts [9]. This kind of network not only achieves the flexible, causal and continuous-time feature of ODE-based networks but also has a better efficiency compared to them [9]. The CfC model can be represented by the equation 3, where *σ* (−*f* (*x, I*; *θ*_*f*_)*t*) and [1 − *σ*(− [*f* (*x, I*; *θ*_*f*_)]*t*)] are the time-continuous gating [9].

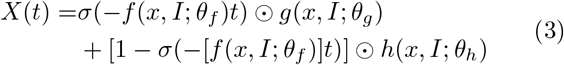

## 3 Datasets

In this study, the proposed models were evaluated using two distinct datasets. The first dataset, referred to as the CPSC dataset or the 12-lead ECG dataset, was created for The China Physiological Signal Challenge 2018 [10]. Its purpose was to facilitate the automatic detection of abnormalities in rhythm and morphology within 12-lead ECGs. The second dataset employed in this research is the Telehealth Network Minas Gerais (TNMG) dataset [11], primarily used for model training.

To assess the models’ performance on new and unseen data, the CPSC dataset was utilized as an independent test dataset. This evaluation aimed to measure the models’ ability to handle real-world data that differs from the TNMG dataset.

The study’s objective was to evaluate the models’ generalization and performance on unfamiliar data by training them on the TNMG dataset and evaluating their effectiveness on the CPSC dataset. This evaluation is crucial for determining the models’ reliability and efficacy in real-life scenarios.

### 3.1 TNMG

The TNMG dataset used in this study consists of 2,322,513 labeled samples of 12-lead electrocardiogram (ECG) data. These samples represent six distinct types of abnormalities: Atrial Fibrillation (AF), First Degree Atrioventricular Block (1dAVb), Left Bundle Branch Block (LBBB), Right Bundle Branch Block (RBBB), Sinus Bradycardia (SB), and Sinus Tachycardia (ST) [11]. The ECG data was originally sampled at 400 Hz frequency.

To create a balanced dataset for model training, 3000 data were selected for each of the six abnormalities at random, along with a further 3000 data without any abnormalities. This resulted in a total sampled dataset size of 21,000. In cases where patients exhibited multiple abnormalities, any remaining samples needed to achieve the subset size of 21,000 were chosen from the TNMG at ramdom. For more detailed information about these six abnormalities, please refer to Table 1.

**Table 1:**
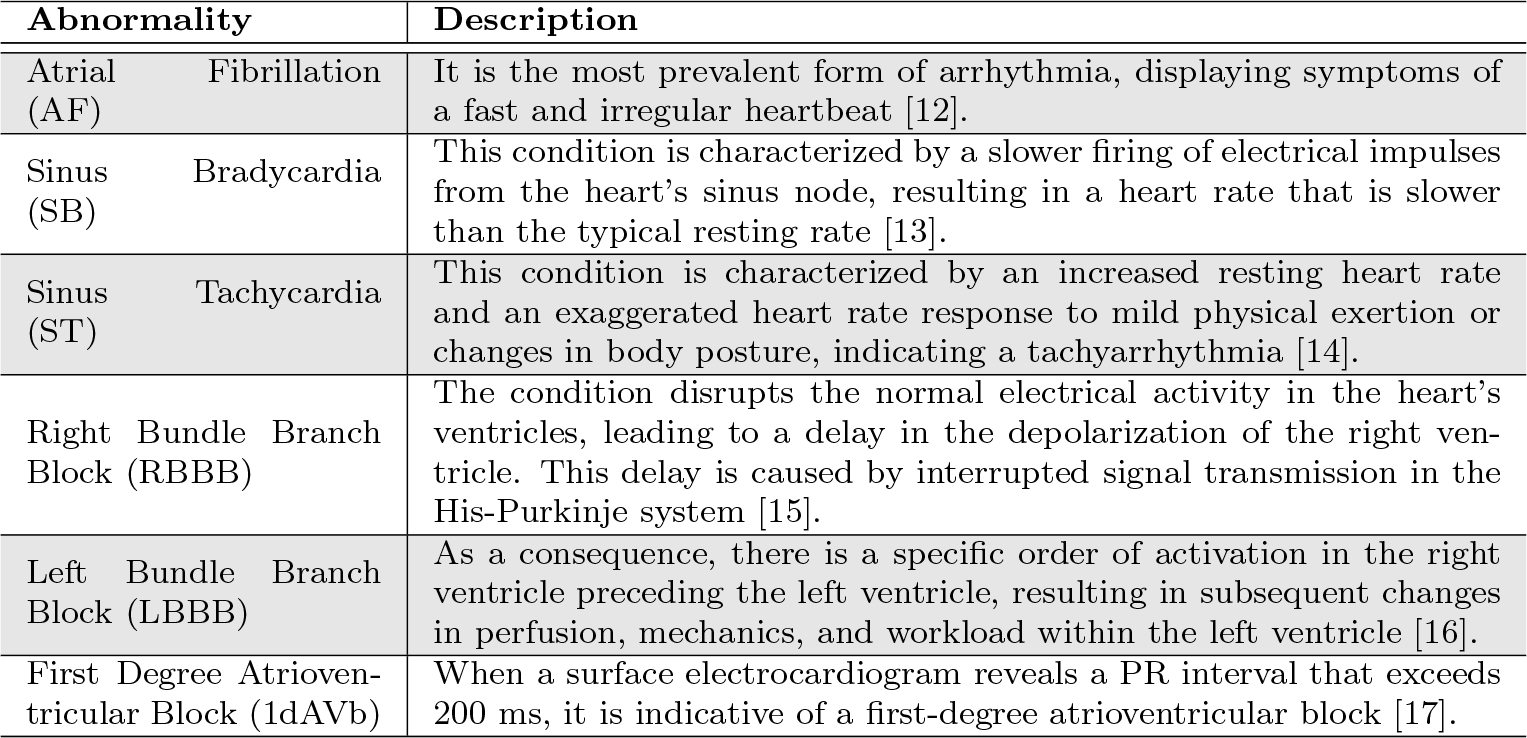
Abnormalities within the TNMG dataset are categorized into different classifications.

The dataset underwent a process of normalization, where it was adjusted to a consistent length of 4096 readings, ensuring uniformity and facilitating analysis and modeling. Any readings exceeding this length were removed, streamlining data processing and comparison. Fig. 2 illustrates a balanced distribution of genders in the resampled dataset, promoting inclusivity and valid analysis. The dataset also reflects the age distribution observed in the general population, enhancing representativeness for age-related analysis. Additionally, the sampling method yielded a balanced distribution of different abnormalities, allowing comprehensive evaluation of their characteristics and impacts. This balanced dataset improves the model’s learning process and overall performance [18].

**Figure 2:**
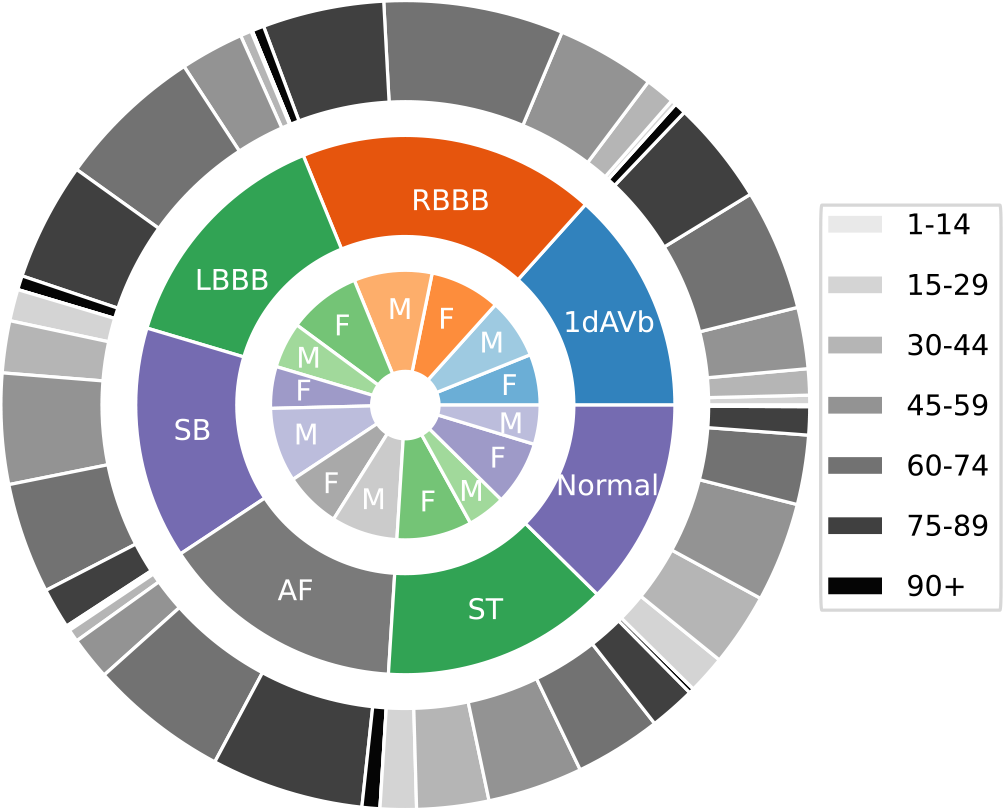
The presented TNMG subset represents a highly balanced dataset containing six types of abnormalities. Additionally, it exhibits a higher concentration of older patients, which is reflective of the general population.

### 3.2 CPSC

The CPSC dataset consists of 12-lead electrocardiograms (ECGs) with a 500 Hz sample rate. To ensure compatibility with the CPSC dataset, the TNMG data was resampled at a rate of 500 Hz specifically for training purposes. This dataset is notable due to its inclusion of electrocardiograms (ECGs) from patients who have been diagnosed with a range of cardiovascular conditions and exhibit common rhythms. The ECGs in the dataset have been expertly labeled, providing accurate annotations for these abnormalities. Overall, the dataset encompasses eight distinct types of abnormalities.

To effectively test the model’s generalization, we conducted tests using four selected abnormalities from the dataset: Atrial Fibrillation (AF), Left Bundle Branch Block (LBBB), Right Bundle Branch Block (RBBB), and First Degree Atrioventricular Block (1dAVb). However, it is important to note that this study does not include four other types of abnormalities: Premature Atrial Contraction (PAC), Premature Ventricular Contraction (PVC), ST-segment Depression (STD), and ST-segment Elevated (STE).

As part of the data selection process, any entries in the dataset with missing readings were excluded, resulting in a final dataset of 6,877 distinct ECG tracings. The data was subsequently standardized by adjusting it to a consistent length of 4,096 readings. Any additional readings beyond this length were removed from the dataset during the cleaning process. For more detailed information about the refined dataset, please refer to Fig. 3.

**Figure 3:**
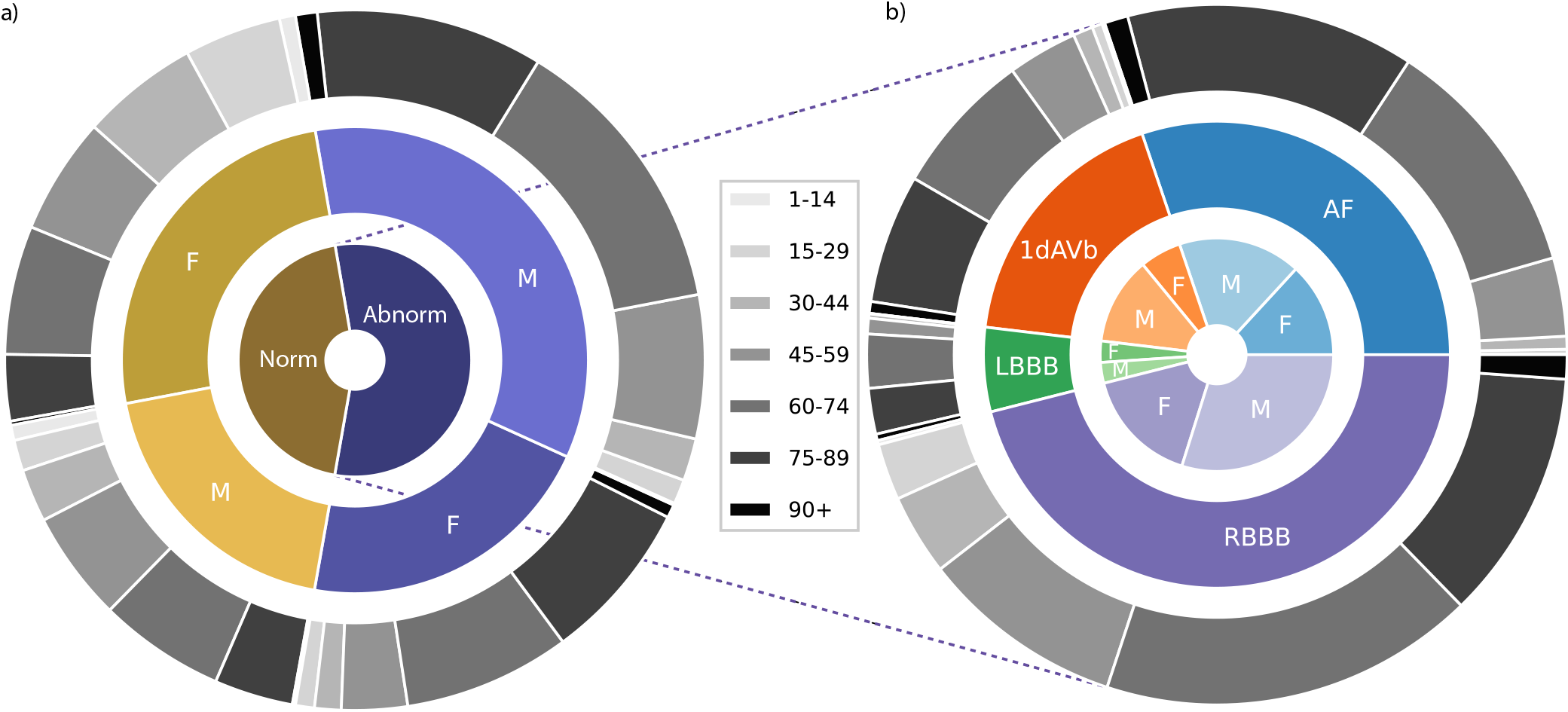
a) plot illustrates the distribution of patients with studied abnormalities versus those without studies abnormalities or with normal ECG readings. b) performs a detailed analysis of patients with studied abnormalities, breaking down the data into specific individual abnormalities.

The analysis of the dataset uncovered a gender disparity, indicating that there is a greater representation of male patients compared to female patients. However, the age distribution of the patients in the dataset aligns with that of the general population, with a larger proportion of individuals belonging to older age groups. However, when examining the distribution of abnormalities, a slight imbalance is observed. Specifically, the occurrence of LBBB is relatively lower compared to the prevalence of other abnormalities present in the dataset.

## 4 Method

Our primary objective is to create a model that is compact, efficient, and well-suited for processing electrocardiogram (ECG) data. We aim to optimize the model specifically for hardware implementation, ensuring its compatibility with resource-constrained devices. Furthermore, we conduct a comprehensive evaluation of the model’s performance in terms of both generalization and robustness to incomplete data.

To achieve our goal, we developed a small-scale architecture that minimizes computational requirements without compromising accuracy. The ECG data is subjected to essential preprocessing steps, such as filtering and short-time Fourier transform (STFT) transformation, to enhance its quality and extract relevant features. Afterwards, the preprocessed data is inputted into the proposed models for the purposes of training and validation. The performance of the model is then assessed using selected performance metrics in order to gain insights into its effectiveness.

### 4.1 Preprocessing of ECG data

To reduce noise interference in the ECG signal, a Butterworth band-pass filter is applied. The Butterworth filter is selected for its uniform response to all desired frequencies [19]. The passband of the band-pass filter is set from 0.5 Hz to 40 Hz. This frequency range is chosen to retain important information such as the T wave, P wave, and QRS complex, while effectively removing powerline noise at 50 Hz [20].

After the signal is filtered, the short-time Fourier transform (STFT) is applied. Unlike the fast Fourier transform (FFT), which operates on the entire signal at once, the STFT divides the signal into smaller windows and applies the FFT to each of these windows individually. This approach allows for the extraction of both frequency and time-related information from the analysis. By analyzing the signal in smaller time segments, the STFT captures changes in the signal over time, providing a more detailed representation of the signal’s time-varying frequency components. This enables the gathering of both frequency and temporal information from the signal analysis. The equation for FFT is shown in equation 4 where X[k] represents complex spectrum at frequency index k, x[n] represents the input sequence of length N and e^−*j*2*πkn/N*^ represents the complex exponential at frequency index k and time index n. The formula for STFT is shown in equation 5, where *X*(*t, ω*) represents the complex value of the STFT at time t, frequency *ω, x*(*τ*) represents the input signal and *w*(*τ* − *t*) denotes the window function centred at time t

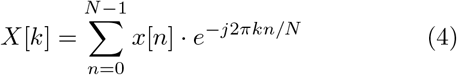

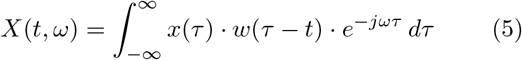

### 4.2 Neural Circuit Policies (NCP)

Inspired by the Caenorhabditis elegans nematode, the neural circuit policies (NCPs) have been developed as a brain-inspired intelligent agent. Unlike contemporary deep models, each neuron within the NCP framework exhibits heightened computational capabilities. Through extensive research, it has been demonstrated that the adoption of NCPs leads to the creation of sparse networks, which, in turn, offer enhanced interpretability compared to conventional models [2].

A NCP network consists of a collection of Liquid Time Constant (LTC) neurons [8]. By implementing the aforementioned NCP design principles, the outcome is highly condensed and sparsely interconnected networks of LTC neurons. LTC neurons in the simulation are commonly represented as leaky integrators, where they accumulate incoming inputs over time and gradually release a portion of the accumulated charge. This leakage mechanism is vital for avoiding saturation of the neuron’s membrane potential and facilitating the processing of temporal information [2].

In addition, this study introduces an independent model for Closed-form continuous-time neural networks (CfC) in the context of time-series modeling. Derived from liquid networks, CfC models outperform advanced recurrent neural networks. They employ closed-form ordinary differential equations (ODEs) and approximate the solution for a previously unsolved integral in liquid time-constant dynamics [9].

### 4.3 Simple Model Architecture

The proposed model architecture, illustrated in Fig. 4, is designed to be simple yet effective. It consists of a single ConvLSTM2D layer responsible for feature extraction, connected to 75 neurons serving as the input neurons for the NCP network. The network can be constructed using either the LTC or CfC arrangement, resulting in two distinct models: CLTC and CCfC models with 14 inter and command neurons and 6 output neurons.

**Figure 4:**
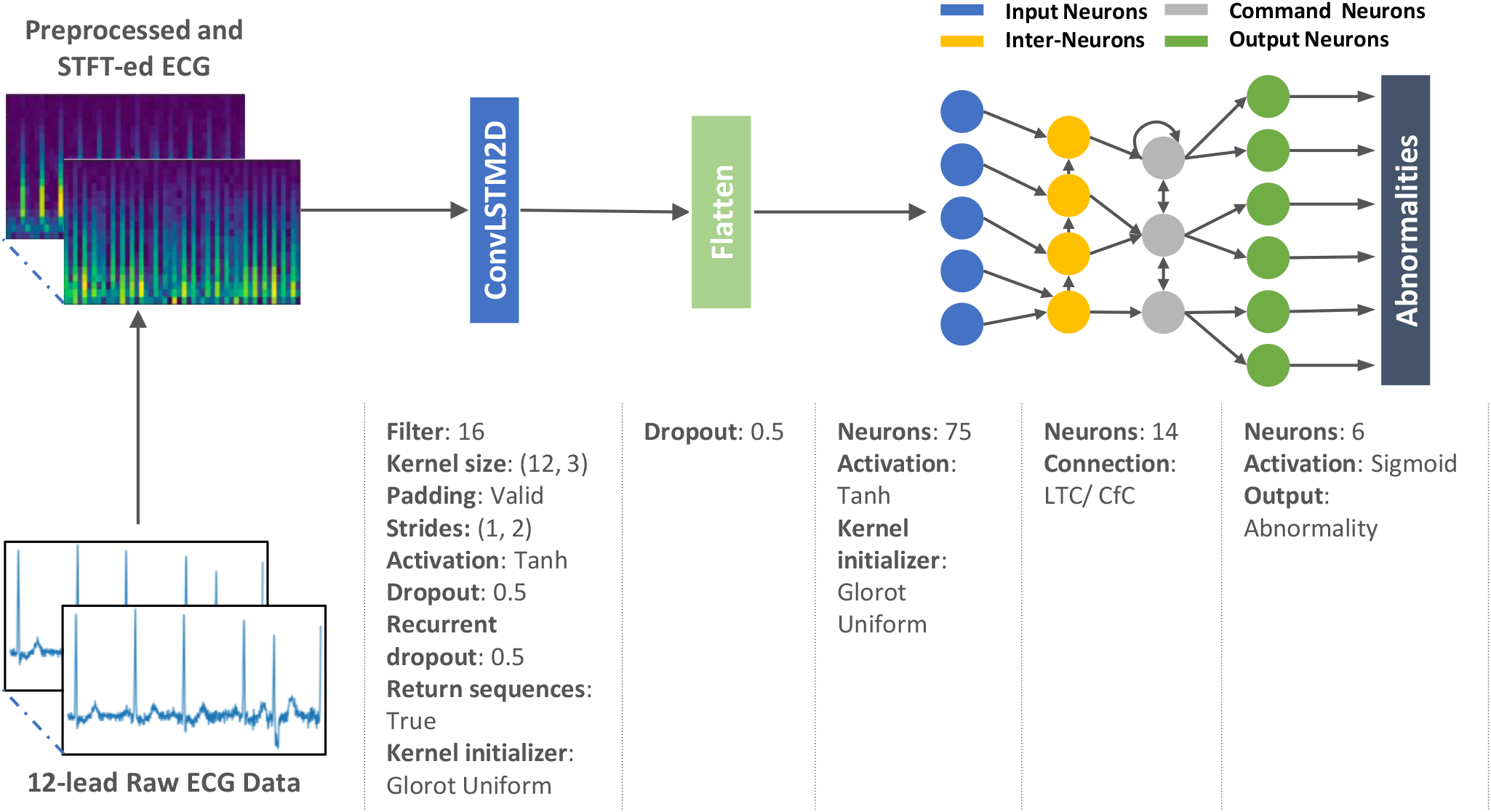
The model architecture consists of a ConvLSTM2D layer that handles a preprocessed ECG data after undergoing STFT transformation [21]. The ConvLSTM2D layer extracts features, which are then densely connected to 75 neurons, serving as the input neurons for the NCP network. The input, motor, and output neurons are interconnected using either LTC or CfC arrangements, with a sigmoid activation function applied at the final stage.

### 4.4 Evaluation Metrics

Precision 7 and recall 6 are metrics used to evaluate the performance of a model, with precision measuring the accuracy of positive predictions and recall capturing the proportion of actual positives correctly identified. The F1-score 8, is the harmonic mean of precision and recall, balancing the precision-recall trade-off.

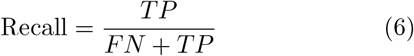

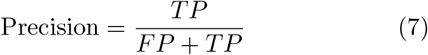

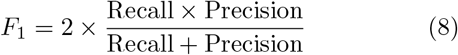

The Area Under the Receiver Operating Characteristic curve (AUROC) is a performance metric employed to evaluate the discriminative capacity of a model in distinguishing between negative and positive cases at different threshold values. It quantifies the model’s ability to classify instances correctly and is commonly employed in binary classification tasks.

These metrics, such as F1-score, precision, recall, and AUROC, are widely used to evaluate the performance of machine learning models in binary classification tasks, particularly in abnormality detection.

## 5 Experiment

### 5.1 Training

In the experimental phase, the preprocessed TNMG subset data is employed as the training dataset, where training and validation procedures are carried out on both the CLTC and CCfC models. To address the class imbalance issue in multi-labeled datasets, a strategy was employed where data instances with positive labels were duplicated twice. This replication technique was implemented to ensure an adequate number of positive-labeled data points within the training dataset, helping to balance the distribution between positive and negative labels. Furthermore, the trained models are deployed onto a microcontroller to showcase their feasibility for chip deployment.

The models were trained on Intel(R) Xeon(R) CPU E5-2630 v3 @ 2.40GHz. During the training process, a batch size 128 was used, and the models underwent 300 epochs of training. The learning rate was set to 0.01, and the Adam optimizer was employed with the binary cross-entropy loss function.

The preprocessed CPSC dataset is utilised to assess the generalization capabilities of the models. Additionally, the robustness of the models is evaluated by testing their performance when exposed to corrupted data inputs.

### 5.2 In-sample Training and Validation

To evaluate the performance of the models, a dedicated subset comprising 20% of the TNMG data was set aside for validation purposes. The specific data points used for evaluation were deliberately withheld from the model’s training process and reserved specifically for assessing the models’ performance. This validation process was conducted independently for each model, enabling a direct comparison of their respective performances.

### 5.3 Model Deployment

After training, the models are deployed onto a microcontroller, and the validation data is utilized to perform inference on the microcontroller. This enables the observation of the deployed models’ performance in a real-world setting, specifically on the microcontroller platform.

### 5.4 Test on Unseen Data

The performance of the models will be thoroughly assessed on the CPSC dataset, which is carefully selected to reflect real-world scenarios. By comparing the models’ predictions with known values using performance metrics, their strengths and limitations will be evaluated. The findings will guide decisions regarding the effectiveness of the models and suggest potential enhancements for better ECG analysis.

### 5.5 Model Robustness

The model will undergo robustness testing by randomly removing channels from the 12-lead ECG data to test its performance when faced with missing inputs. The tests will progressively remove different numbers of channels (1 to 6 leads) to evaluate the model’s ability to maintain accuracy. Performance metrics will be employed to assess and compare how well the model performs in various scenarios. These metrics will help identify areas where the model may need improvement and provide insights for potential enhancements.

## 6 Result

This section will provide a comprehensive analysis of the models’ performance. Moreover, the discussion will also include an in-depth exploration of their generative ability and robustness.

### 6.1 Performance of the Models

This research study primarily revolves around training the proposed models using the TNMG subset dataset, as previously mentioned in the paper. The TNMG subset dataset serves as the main training data for developing and evaluating the models in this study. The training process was conducted over 300 epochs, with careful monitoring and recording of metrics such as accuracy and loss for both the validation and training sets at each epoch. The figures presented in the paper visually illustrate the recorded metrics, providing a comprehensive overview of the entire training procedure. Notably, the performances of both the CLTC and CCfC models were assessed using identical settings, and their individual performances are plotted in Fig 5.

**Figure 5:**
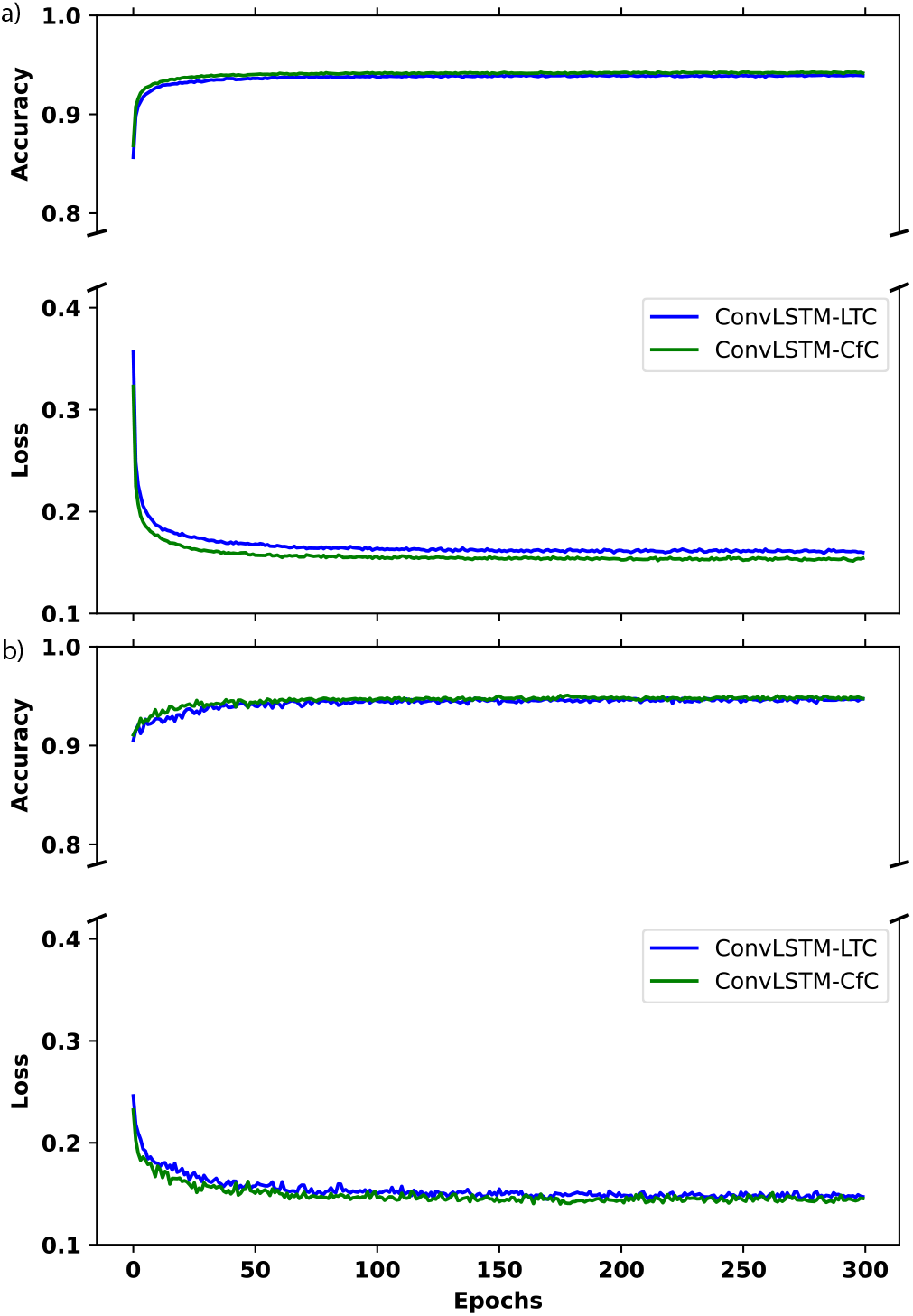
a) The figure visualizes the performance of the two models during the training process. b) The figure showcases the performance of loss and accuracy metrics in validation.

Fig. 5a) demonstrates a consistent decrease in training loss as the training progresses, indicating that the model’s performance improves over time. Simultaneously, the training accuracy steadily increases and eventually reaches a stable level, indicating that the model successfully learns from the training data. Although both models have achieved similar results, the CCfC model has slightly higher accuracy than the CLTC model.

Throughout the training process, a general upward trend is observed in the validation accuracy, indicating an improvement in the model’s performance on unseen data. Additionally, the validation loss consistently decreases over time, suggesting that the model’s predictions align more closely with the ground truth labels during validation. Additionally, Fig. 5b) illustrates that the two models behave similarly, but the CCfC model continues to outperform the CLTC model slightly.

The performance of the CLTC and CCfC models is summarized in Table 2 and Table 3 respectively. Both models have achieved comparable performance, with an F1 score of 0.827 for CLTC and 0.828 for CCfC and AUROC values of 0.961 for CLTC and 0.963 for CCfC. These performance metrics align with the trends observed in Fig. 5. In addition to differences in performance and accuracy, another notable observation between the two models is their contrasting training speeds. The CCfC model demonstrates faster training compared to the CLTC model. Specifically, the CCfC model trains approximately 40% faster than its CLTC counterpart. It’s worth noting that both models were trained on CPUs instead of GPUs. This observation suggests that the simple model architecture requires fewer calculations and less computational power during training.

**Table 2:** CLTC Model Validation Results (P: Precision, R: Recall)

| Class | P | R | F1 | AUROC |
| --- | --- | --- | --- | --- |
| 1dAVb | 71.2% | 52.7% | 60.6% | 88.1% |
| RBBB | 92.5% | 84.0% | 88.1% | 97.2% |
| LBBB | 91.2% | 93.6% | 92.4% | 99.1% |
| SB | 89.3% | 92.6% | 90.9% | 99.0% |
| AF | 77.5% | 70.5% | 73.8% | 94.1% |
| ST | 92.5% | 88.7% | 90.6% | 99.3% |
| Average | 85.7% | 80.3% | 82.7% | 96.1% |

**Table 3:** CCfC Model Validation Results (P: Precision, R: Recall)

| Class | P | R | F1 | AUROC |
| --- | --- | --- | --- | --- |
| 1dAVb | 75.9% | 52.2% | 61.8% | 89.3% |
| RBBB | 92.3% | 82.9% | 87.3% | 97.1% |
| LBBB | 88.9% | 93.2% | 91.0% | 99.1% |
| SB | 89.2% | 92.9% | 91.0% | 99.0% |
| AF | 78.7% | 71.3% | 74.8% | 93.7% |
| ST | 90.4% | 91.5% | 90.9% | 99.3% |
| Average | 85.9% | 80.6% | 82.8% | 96.3% |

### 6.2 Model Deployments on Micro-controller

This experiment successfully deployed CLTC and CCfC models on an STM32F746G Discovery board, a resource-constrained edge device featuring an STM32F746NGH6 Arm Cortex core-based microcontroller. With its 1 Mb of flash memory and 340 Kbytes of RAM, this board proved capable of managing the complex computational tasks required for neural network inference. The software solution STM32Cube.AI, developed by STMicroelectronics, was instrumental in optimising the conversion and deployment of the pre-trained models for the STM32 microcontrollers.

Both models were compared to the previous batch of 30 inferences executed on the STM32F746G Discovery board. It reported a precision of 0.88, and the recall was approximately 0.72. The F1 score, a measure that balances precision and recall, was approximately 0.79, indicating well-rounded model performance.

Similarly, the CCfC model also displayed notable performance. The model reported a commendable precision of 0.97. However, the recall was about 0.66, slightly lower than the CLTC model. The F1 score was approximately 0.76, illustrating a fairly balanced model performance regarding precision and recall.

When the interface runs at a clock frequency of 200 MHz, the power consumption is measured at 668.7 mW. On the other hand, when the interface is not running, the power consumption is observed to be 531.3 mW. As a result, the model itself consumes approximately 137.4 mW. These results represent a significant milestone for machine learning in edge devices, showcasing the viability of deploying sophisticated deep learning models on resource-constrained hardware. However, there remains ample room for improvement. Future work will explore the potential of deploying the models using TinyEngine, a compact inference library developed by MIT researchers [22, 23]. This platform may enhance the accuracy of the neural network model when inference is performed on edge devices, further advancing AI on edge devices [22, 23]. The findings of this experiment highlight the feasibility and potential of deploying deep learning models in such environments, opening a new frontier for AI applications.

### 6.3 Generalization of Models

Predictions were made on the CPSC dataset using the trained models on the TNMG subset, as described in the data section, to assess the generalisation ability. This evaluation aimed to measure how effectively the models could handle new and unseen data from the CPSC dataset, which differs from the training dataset. It is important to mention that the CPSC dataset contains eight different types of abnormalities, but only four of them are also present in the TNMG dataset. By evaluating the chosen model’s performance on this subset, we can gather valuable information about its capacity to apply learned knowledge to new and unseen data.

The performance of the two models on unseen data is presented in Table 4 and Table 5, demonstrating their ability to perform well. The CLTC model achieves an average F1 score of 0.70 and an AUROC of 0.90, while the CCfC model achieves an F1 score of 0.72 and an AUROC of 0.91. These results show that both models possess strong generalization capabilities, with the CCfC model exhibiting slightly better generalization performance than the CLTC model.

**Table 4:** CLTC Model Generalization Results (P: Precision, R: Recall)

| Class | P | R | F1 | AUROC |
| --- | --- | --- | --- | --- |
| 1dAVb | 47.8% | 65.2% | 55.2% | 86.7% |
| RBBB | 91.5% | 60.8% | 73.0% | 83.8% |
| LBBB | 89.0% | 82.6% | 85.7% | 96.6% |
| AF | 50.8% | 92.9% | 65.7% | 93.4% |
| Average | 69.8% | 75.4% | 69.9% | 90.1% |

**Table 5:**
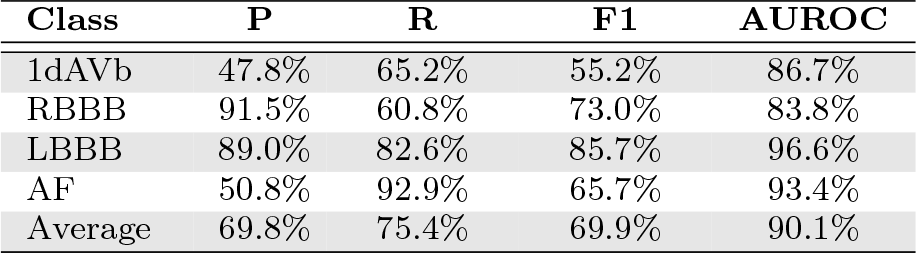
CCfC Model Generalization Results (P: Precision, R: Recall)

### 6.4 Model Robustness

To further assess the model’s performance, we conducted additional evaluations on the CPSC dataset by randomly removing certain channels from the 12-lead ECG data. This allowed us to investigate the model’s robustness and capability to handle incomplete or missing input information.

In Fig. 6, it can be observed that as the number of emptied leads increases, the F1 performance metric of the models declines. Initially, the CCfC model demonstrates superior performance compared to the CLTC model.

**Figure 6:**
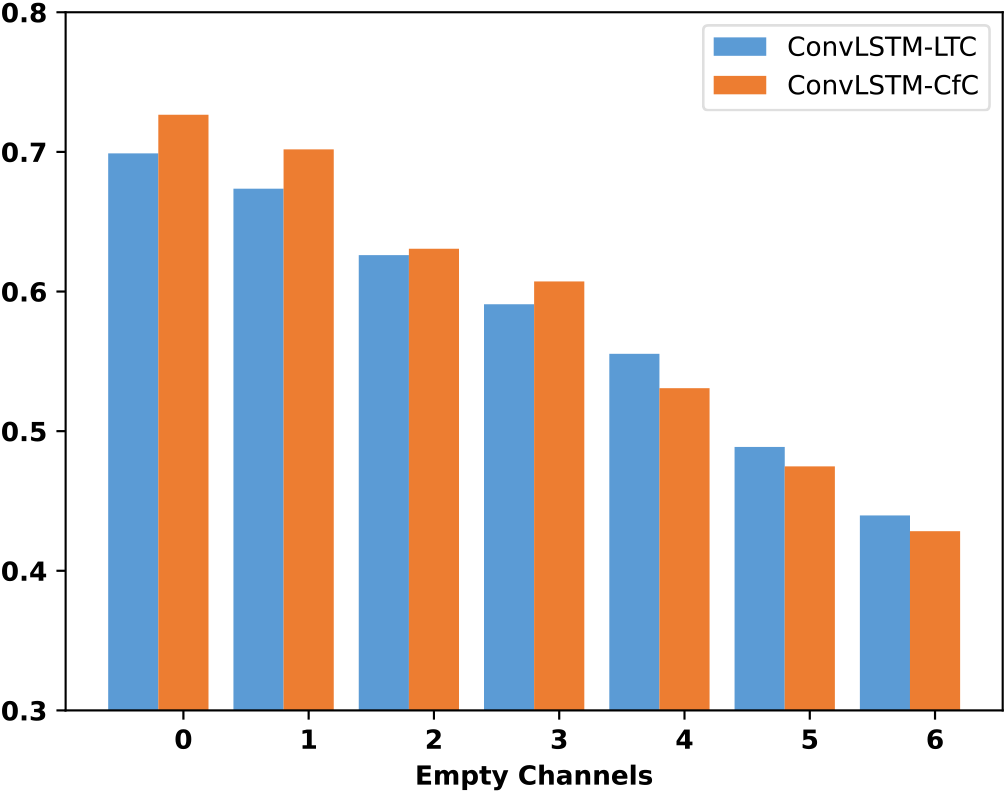
The F1 metrics of the models were evaluated based on the number of empty channels in the 12-lead CPSC ECG data.

However, an interesting finding emerges as the number of empty channels increases: the CLTC model starts to outperform the CCfC model. This suggests that the CLTC model may exhibit better robustness when faced with more empty channels.

## 7 Discussion

The results provide a comprehensive analysis of the models’ performance, their generative ability, and robustness.

Regarding the performance of the models, both the CLTC and CCfC models achieved similar results in terms of F1 scores and AUROC values. The CCfC model demonstrated slightly higher accuracy compared to the CLTC model but with an increasing number of empty channels, the CLTC model showed a higher accuracy, which means the CLTC model had a better robustness when faced with a higher number of empty channels compared to the CCfC model. The training process showed that the models consistently improved their performance during training, with decreasing loss and increasing accuracy. The validation results also indicated that both models performed well, with the CCfC model slightly outperforming the CLTC model.

The models were successfully deployed on a resource-constrained microcontroller, the STM32F746G Discovery board, demonstrating the feasibility of running complex deep learning models on edge devices. The deployed models achieved commendable precision and recall, resulting in the CCfC model exhibiting faster training speed than the CLTC model.

The generalization performance of the models was evaluated on the CPSC dataset, which included abnormalities not present in the training data. Both models demonstrated good generalization capabilities, with the CCfC model showing slightly better performance regarding F1 scores and AUROC values.

Overall, the results demonstrate the effectiveness and potential of the proposed models for abnormality identification. The models showed promising performance, generalization capabilities, and the ability to handle incomplete or missing input information. Further improvements can be explored, such as deploying the models with optimized inference libraries for edge devices, which may enhance their accuracy in resource-constrained environments.

## 8 Conclusion

In conclusion, using ECG data, this study demonstrated two models, CLTC and CCfC, for abnormality identification. Both models demonstrated comparable performance and good generalization capabilities on unseen data. The models were successfully deployed on a resource-constrained microcontroller, showcasing their potential for edge device applications. The findings highlight the effectiveness of the models in abnormality detection and their ability to handle incomplete input. Further improvements can be explored, such as optimizing inference libraries and expanding the training data. Overall, these models contribute to the advancement of AI in healthcare and hold promise for early detection and treatment of cardiac conditions.

## Data Availability

This research paper utilizes the publicly accessible CPSC dataset for analysis and experimentation. However, it is crucial to acknowledge that the TNMG dataset, on the
other hand, is not publicly available. To gain access to the TNMG dataset, permission must be obtained from the data owner. Although access can be granted upon approval, it is important to note that the TNMG dataset is not freely accessible to the general public.

## 9 Acknowledgement

Zhaojing Huang expresses gratitude for the Australian Government’s Research Training Program (RTP) support.

Luis Fernando Herbozo Contreras acknowledges the partial support from the University of Sydney in the form of the Faculty of Engineering Research Scholarship.

## 10 Availability of Code

Share your requests with the corresponding author to access the code used in this research paper. It is important to note that there might be specific terms and conditions or restrictions on the usage of the code, which will be communicated to you by the author.

## 11 Availability of Data

This research paper utilizes the publicly accessible CPSC dataset for analysis and experimentation. However, it is crucial to acknowledge that the TNMG dataset, on the other hand, is not publicly available. To gain access to the TNMG dataset, permission must be obtained from the data owner. Although access can be granted upon approval, it is important to note that the TNMG dataset is not freely accessible to the general public.

## 12 Conflict of Interest Statement

The authors declare that they have no conflicts of interest, including both financial and non-financial, to report.

## Notes

### Competing Interest Statement

The authors have declared no competing interest.

### Author Declarations

CPSC dataset is a publicly available ECG dataset, while TNMG ECG dataset is a private dataset requested from the author of the article 'Automatic diagnosis of the 12-lead ECG using a deep neural network'.

